# Impressions of Vibrotactile Signals in Older Adults With and Without History of Stroke

**DOI:** 10.1101/2020.08.21.20179432

**Authors:** Caitlyn E. Seim, Brandon Ritter, Kara E. Flavin, Maarten G. Lansberg, Allison M. Okamura

## Abstract

Vibrotactile feedback is mechanical stimulation produced using actuators in contact with the body. The stimulation parameters (frequency, amplitude, location, duration) can be adjusted to produce a variety of sensations. By characterizing how users respond to different settings, interaction designers can create more usable and enjoyable haptic interfaces. This form of haptic feedback is being used widely for alerts, gaming, and simulation; however, emerging technologies in the fields of brain health and physical therapy are introducing new users to this stimulation. For applications using vibrotactile stimulation to advance, researchers are studying perceived sensations and affective response. However, these studies often focus on healthy, younger users. It is well known that older adults and those with acquired brain injury have different physiology and different perception than young adults. Here we present a set of vibrotactile signals to adults over 40 years old with and without history of stroke and query affective impression and experienced sensations. Signals on the palm and those with a changing stimulus location were associated with higher valence ratings, while low-amplitude signals showed lowest arousal. Users preferred stimulation that they could perceive, and they could not perceive most signals applied to the forearm. Reported sensations include tickle, tingling, and numbness.

**CCS Concepts:** Human-centered computing → User studies; Haptic devices; Ubiquitous and mobile devices.

## 1 Introduction

Vibrotactile stimulation is an unobtrusive form of haptic feedback commonly integrated into mobile, wearable and interactive devices. Vibrotactile stimulation is being used widely for alerts, gaming and simulation; but lately physical therapy is an emerging application that attracts new user demographics. To improve interactions using vibrotactile stimulation, researchers are studying affective response and user experience at various stimulation settings. These studies often focus on healthy, younger users; however, older users of new health and therapeutic applications will have different sensory perception than these younger users. In the interest of developing inclusive technology that impacts the aging population, the voice of these prospective users should be integrated into the design process. Here we present a variety of vibrotactile signals to older adults with and without history of stroke, and query their affective impression and experienced sensations in response to these signals. The contributions of this paper are: (1) affective response to stimulation at various settings from adults over 40 with and without history of stroke, (2) themes of user preferences for stimulation, (3) reported sensations during hand and arm vibration.

## 2 Background and Motivation

Vibrotactile feedback is mechanical stimulation produced using actuators in contact with the body. The stimulation parameters (frequency, amplitude, location, duration) can be adjusted by system designers to produce a different user experience. Stimulation may induce different feelings in the body, such as tingling, or imitate tactile interactions such as a button click [Markow et al. 2010]. In addition to sensations, users can ascribe affective impressions to signals, for example, finding some stimuli more arousing and energetic [Seifi and Maclean 2013; Yoo et al. 2015]. By characterizing how users respond to different settings, interaction designers can create more usable and enjoyable haptic interfaces.

Vibrotactile stimulation is already pervasive, with use in cell phones [Han et al. 2014], gaming [Shirali-Shahreza and Shirali-Shahreza 2009], surgery [Petter et al. 1996] and training [Eriksson et al. 2006] for example. Human perception and application-specific use of haptic feedback is well studied, but these evaluations do not focus on affective response and preferences. To improve the design of such systems, and expand the application space, researchers are now evaluating human subjective response to stimuli. For example, Seifi et. al. examined lifestyle differences as having a potential role in user experience of vibrotactile sensations [Seifi and Maclean 2013]. In “Emotional responses of tactile icons,” Yoo et. al. used valence and arousal to evaluate tactons varied by amplitude, frequency, duration, and envelope [Yoo et al. 2015]. They found higher frequencies to have higher valence, and higher amplitudes to have higher arousal. Hasegawa et. al. performed a similar evaluation and found similar results in terms of valence and arousal [Hasegawa et al. 2019]. Culbertson et. al. used very low frequency, lateral activation patterns and found longer activations with overlapping activation times most pleasant [Culbertson et al. 2018]. Nunez et. al. evaluated affective response using rotating tactors and found speed and delay were factors in pleasantness ratings [Nunez et al. 2019]. Further, Knibbe et. al. performed a similar evaluation of user experience of electrical stimulation [Knibbe et al. 2018]. However, such work often focuses on healthy, adult participants under 40 (18–30 years old [Culbertson et al. 2018], 18–31 [Yoo et al. 2015], 24–32 [Wilson and Brewster 2017], 21–24 [Hasegawa et al. 2019]).

An increasing number of emerging technologies focus on an older demographic. Older users may need a reason to engage with haptic technology, and for many that comes when seeking to improve brain health or undergoing rehabilitation. Physical therapy is one of the main emerging applications for haptic feedback because of its direct interaction with the somatosensory system [Backus et al. 2014; Cordo et al. 2009, 2013; Dinse et al. 2006; Enders et al. 2013; Lakshminarayanan et al. 2015; Markow et al. 2010; Seim 2019; Seo et al. 2014; Sharma et al. 2011]. Vibrotactile stimulation is being used as part of physical therapy in stroke survivors [Cordo et al. 2009, 2013; Seim 2019] as well as other populations [Backus et al. 2014; Markow et al. 2010]. Similarly, vibrotactile stimulation is being applied to restore sensation with age [Dinse et al. 2006; Enders et al. 2013], to enhance motor function during task performance [Seo et al. 2014], and to improve cutaneous sensation [Lakshminarayanan et al. 2015]. Furthermore, applications such as VR games for older users apply stimulation to render feedback [Ijsselsteijn et al. 2007; Kapur et al. 2009; Molina et al. 2014]. In ‘Digital Game Design for Elderly Users’ IJsselsteijn et. al. highlight the same sensory and physical differences in older users that we call to attention here [Ijsselsteijn et al. 2007]. These therapeutic applications attract a demographic of older adults, with and without history of brain injury, to use haptic technologies.

Just like other populations, older users may have preferences for the stimuli and sensations that they encounter. However, older adults are known to have a different response to somatosenory stimulation than younger users who are more often studied by system designers. Somatosensation has been studied in older adults [Cholewiak and Collins 2003; Dinse et al. 2006; Frisina and Gescheider 1977; Kenshalo 1979; Kenshalo Sr 1986; Stevens 1992; Verrillo 1980, 1982, 1993], but these studies do not focus on affective response or user experience. The user-centered design process is key to development of human-facing technologies. Take for example, technology design for differently-abled populations. Including these users in the design process is considered essential to making new devices for them [Ladner 2015]. “User opinion is key to adoption of assistive devices,” said Manns et al. in their 2019 paper [Manns et al. 2019]; while Norman calls attention to users’ emotional response to technologies [Norman 2004]. Similarly, older adults have different sensory perception, beliefs and experiences which inform their interaction with technology.

Here we query adults over 40, with and without history of stroke, to reveal their impressions of vibrotactile stimuli. We use the same validated metrics used in prior work to collect the affective response of our participants. Our first aim is to examine whether any signals are considered noxious or intolerable by our participants. We also aim to assess user experiences of each stimulation – to inform the development of more user-conscious systems.

### 2.1 Somatosensation in Older Adults

Tactile perception and sensation change with age. Starting in their 20s, individuals gradually lose sensitivity to touch, including pressure and vibratory stimulus [Cholewiak and Collins 2003; Dinse et al. 2006; Frisina and Gescheider 1977; Kenshalo 1979; Kenshalo Sr 1986; Stevens 1992; Verrillo 1980, 1982, 1993]. These reductions in perceptive ability have been attributed to both changes in skin and changes in the central nervous system that occur with age. Sensory receptors within the skin change in number and structure as a person ages [Besné et al. 2002; Montagna 1965; Steinberg and Graber 1963; Stevens 1992; Verrillo 1979], including the Meissner and Pacinian corpuscles which are most responsive to vibration [Goble et al. 1996; Schimrigk and Rüttinger 1980; Winkelmann 1965]. Changes in the sensorimotor brain map areas [Kalisch et al. 2008], and conditions such as neuralgia or diabetes are also found in older adults to relate to tactile perception [Lundström and Lindmark 1982; Mirsky et al. 1953; Wahren and Torebjörk 1992].

### 2.2 Stroke and Perception

Nearly three-quarters of all strokes occur in people over the age of 65 and the risk of having a stroke more than doubles each decade after the age of 55. Stroke occurs when part of the brain cannot get enough oxygen, typically resulting from a blood clot or hemorrhage. Stroke survivors are left with a damaged region of the brain, often in the sensorimotor area which controls movement and sensation – leading to chronic physical disability [Yu et al. 2016].^1^ Over 15 million people have a stroke each year, making it one of the leading causes of disability in the United States and worldwide [Adamson et al. 2004; Bonita et al. 2004; for Disease Control et al. 2012].

Stroke survivors may use stimulation for rehabilitation or may encounter it in other contexts, and these individuals also have abnormal sensory function at a rate of 50–80% due to their brain injury [Connell et al. 2008; Kim and Choi-Kwon 1996]. Survivors may have lower cutaneous tactile perception, making them less sensitive to touch and pressure sensations [Connell et al. 2008]. Some individuals with history of stroke also experience ***increased*** sensitivity to tactile stimulation such as heat, cold, and pin prick [Boivie et al. 1989; Bowsher 2005; Greenspan et al. 2004] and may experience these non-noxious simulations as pain (allodynia). Stroke survivors may also show errors in proprioception and kinesthetic awareness, and may experience sensory disturbances such as neuropathy (tingling nerve pain).

## 3 Apparatus

Three wearable devices were designed to apply stimuli for this experiment. Each device stimulates a different area of the upper limb. Actuator sites are depicted in Figure 2. Participants try each device during the study to obtain an impression of the signals at different locations: the phalanges, the palm, and the forearm. These locations on the body each have different tissue structures and distribution of sensory receptors [Johansson and Vallbo 1979; Lee 2010]. Each device contains five actuators (of each type). The devices are ambidextrous and one-size-fits-all, designed with accommodations to allow stroke survivors to don and doff using only one hand.

**Fig. 1.**
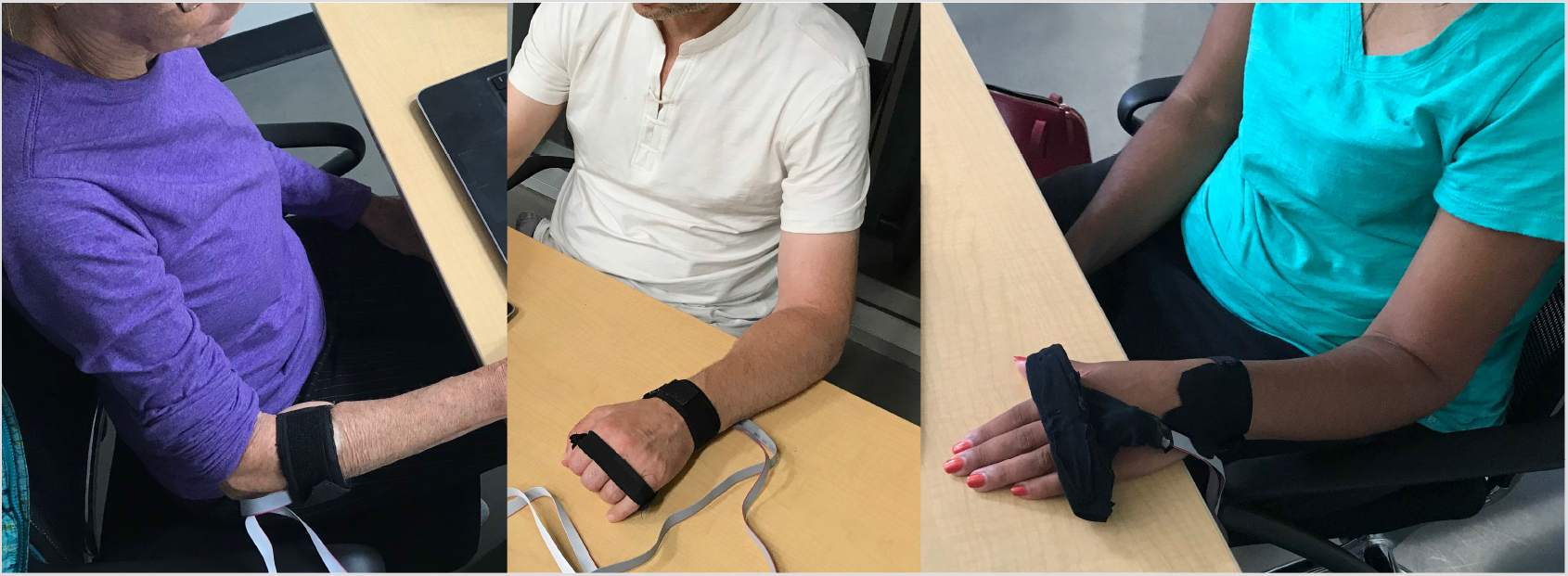
Participants wearing stimulation devices during the study.

**Fig. 2.**
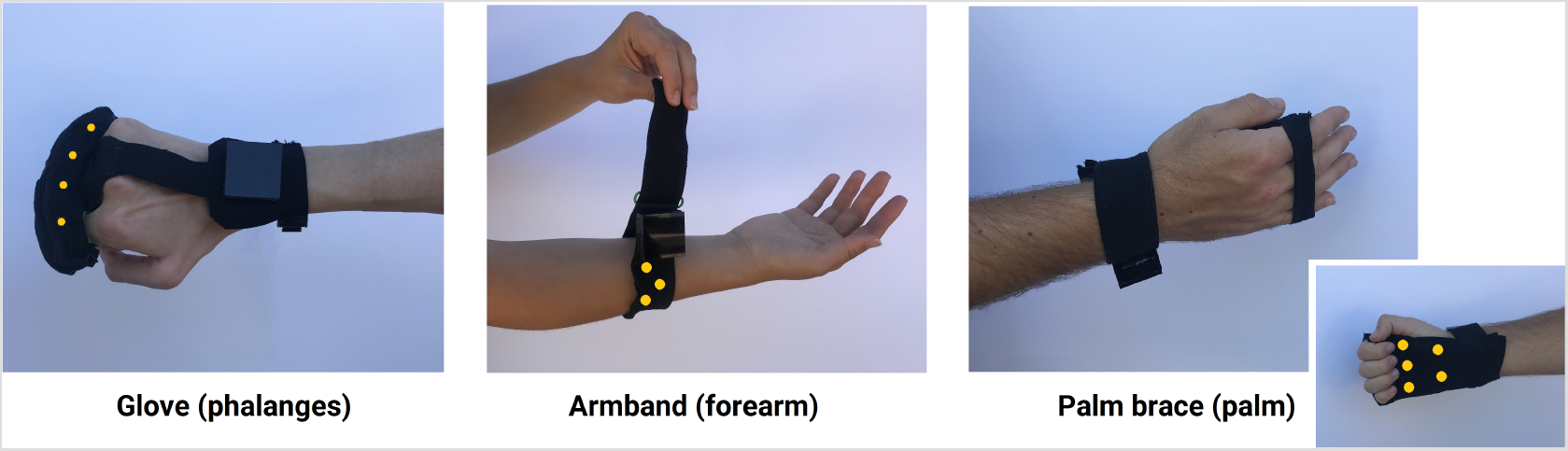
Three stimulation devices used in the study. Each device contains 5 actuators of each type. Yellow dots are added to this figure to indicate actuator locations; not all actuators are visible here.

### 3.1 Stimuli

We aimed to test user impressions of stimuli at a various settings. We designed 12 vibrotactile signals to provoke a variety of responses from the somatosensory system which may be of use various applications of vibrotactile stimulation. Three dimensions are used to create the 12 signals, including actuator settings (frequency and amplitude), pattern (spatial), activation duration (temporal). Figure 3 shows a representation for each signal.

**Fig. 3.**
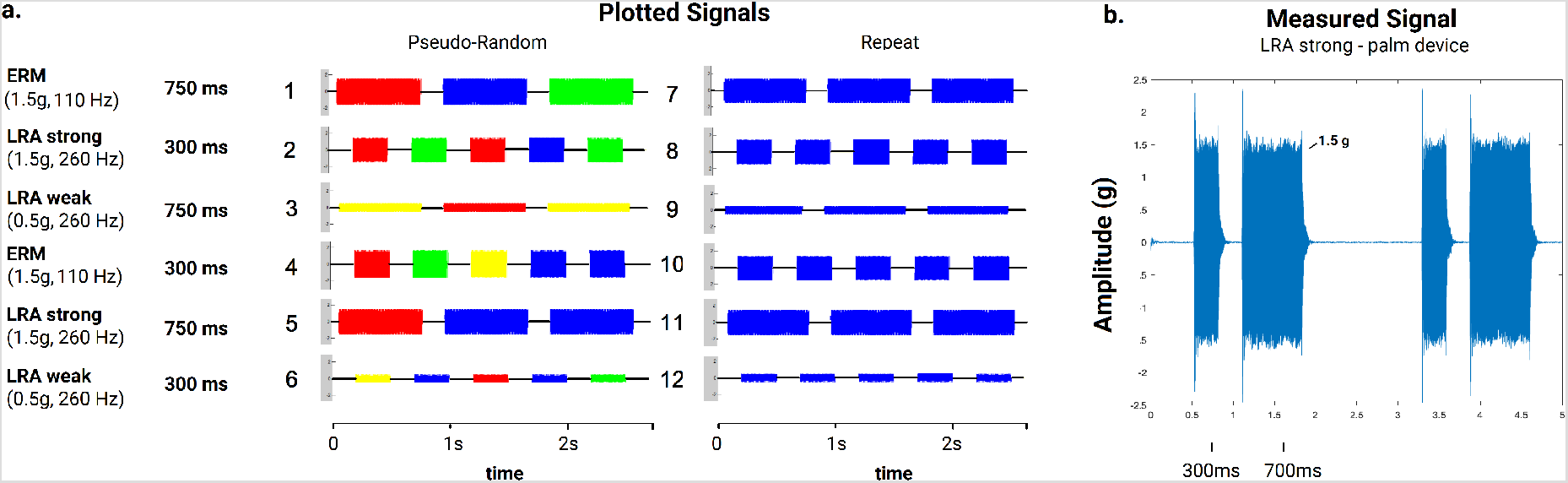
**a.** 2.7 second plot of each stimulus (signals are 30 seconds long in total). Numbers 1–12 are used to refer to each signal. Changing colors indicate changing stimulus location in the pseudo-random patterns. **b.** Example accelerometer measurement of LRA actuator when embedded into the palm device.

#### 3.1.1 Actuators Settings

We test two frequencies in this experiment – one tuned to the Pacinian corpuscles (a cutaneous mechanoreceptor which preferentially responds to high-frequency vibration around 240 Hz [Johansson and Vallbo 1979; Scheibert et al. 2009]) and one tuned to the muscle afferent fibers (sensory organs of the muscles which preferentially respond to vibration around 70–80 Hz [Cordo et al. 1993, 2009]). Stimuli that target different structures may be helpful to system designers and users may notice a difference between these low- and high-frequency settings.

We also test different amplitudes in this experiment. The amplitude and frequencies of stimuli are limited by actuator capabilities. Here we aim to use actuators that could be integrated into a mobile or wearable format.There are limited actuators available to provide vibrotactile stimulation, and fewer that can be integrated into a mobile form factor. While high-frequency stimulation is possible at a range of amplitudes using Linear Resonant Actuators (LRAs), small actuators that provide a range of amplitudes at a frequency below 175 Hz are not available. Here we use an Eccentric Rotating Mass (ERM) motor (Precision Microdrives 310–103) to provide the lower frequency stimulation at around 90–110 Hz. At this frequency, the motor has a fixed amplitude measured at around 1–1.5 g.^2^ For our high-frequency stimulation, we use an LRA motor (Precision Microdrives C08–005) which is capable of providing stimulation at a range of amplitudes at this frequency (measured around 240–260 Hz). We select 0.5 g and 1.5 g as the target amplitudes using this motor. We measured the characteristics of each signal using a Kistler accelerometer (Type 8694M1) affixed to each motor using putty (Quake Hold Museum Wax). Table 1 shows measured stimuli characteristics in each device, as these characteristics change when actuators are embedded into fabric. Since frequency and amplitude are tied to actuator capabilities, here we use the motor names to refer to the three conditions of vibration settings.

**Table 1.** Measured stimuli characteristics for actuators alone (Device – N/A) and in each device. “LRA weak” refers to the low-amplitude LRA stimuli,“LRA strong” refers to the high-amplitude LRA stimuli. No stimuli names are shown to participants, they only sense the stimuli from the devices.

| Device | ERM |  | LRA strong |  | LRA weak |  |
| --- | --- | --- | --- | --- | --- | --- |
| N/A | 1.5 g | 110 Hz | 1.5 g | 259 Hz | .5 g | 260 Hz |
| Glove | 1.5 g | 100 Hz | 1.5 g | 240 Hz | .5 g | 265 Hz |
| Palm | 1.5 g | 90 Hz | 1.5 g | 240 Hz | .5 g | 275 Hz |
| Armband | 1.5 g | 90 Hz | 2 g | 250 Hz | .75 g | 260 Hz |

#### 3.1.2 Pattern

Two spatial patterns are tested in this work. One is a pseudo-random pattern, where stimulus location changes between the five actuators. Each location is activated the same number of times, but in a random order. Figure 3 indicates the changing stimulus location of pseudo-random signals by using multiple colors. The second pattern activates the same location repeatedly – five times in a row, before moving to an adjacent location. These patterns will feel different to users. In addition, this repeated stimulation is designed to be adapting or habituating to the user. Adaptation to a stimulus occurs after repeated or with increasing duration of stimulation to an area, and causes reduced sensitivity to the same stimulus [Tannan et al. 2006]. This pairs with increased sensitivity to stimuli of different characteristics or stimuli in other areas. In contrast, the pseudo-random stimulation is designed to minimize these effects.

#### 3.1.3 Activation Duration

Two temporal settings are used in this work. Based on pilot testing, one is designed to give a short pulse (300 ms) and the other is a long pulse (750 ms). Longer stimulus duration enhances adaptation compared to shorter stimulus [Tannan et al. 2006].

## 4 Methods

Participants visited the lab for one 1.5 hour session. Each person tried all three devices (one at a time) during the study to examine the signals at different locations. The device order was randomized and counterbalanced. Each device was used to present all 12 signals in a random order. A signal was applied for 30 seconds, then the user had one minute to take a survey on their impression and experienced sensations. Then the next stimulus was applied, and so on.

Stroke survivors wore the devices on their affected side (stroke causes unilateral impairment) and the unaffected older adults group were assigned using a list of the side tested in stroke survivors, regardless of handedness. We hypothesized that all participants would prefer low-amplitude stimulation, and that stroke survivors would sense some high-amplitude signals as painful.

### 4.1 Participants

Two cohorts of participants were recruited through community flyers. The study contained twelve participants: stroke survivors (mean age = 53.5 years) and participants of similar age (mean age = 54.2 years) without history of stroke. Their demographic information is shown in Table 2.

**Table 2.** Participant demographics. Stroke survivors are S1-S6, Unaffected users (without history of stroke) are U1-U6. “Side affected” refers to the side most impacted by stroke. Semmes-Weinstein monofilament exam (SWME) scores of tactile acuity are reported on a scale of 1–5 and are averaged over the tested locations. Hand dominance is self-reported.

| User ID | Age | Hand Dominance | Side Affected | Study Side | Average SWME<br>Palm | Average SWME<br>Dorsal Hand | Average SWME<br>Forearm | Notes |
| --- | --- | --- | --- | --- | --- | --- | --- | --- |
| S1 | 50s | R | L | L | 1 | 1.75 | 1.38 |  |
| S2 | 40s | R | R | R | 1 | 1.75 | 2 |  |
| S3 | 80s | R | R | R | 1 | 1 | 1 |  |
| S4 | 40s | R | L | L | 1 | 1.5 | 1 | self-reported allodynia |
| S5 | 50s | R | L | L | 2.5 | 1.25 | 1.38 | self-reported allodynia |
| S6 | 40s | R | L | L | 2 | 1 | 2.5 |  |
| U1 | 60s | R | N/A | R | 1 | 1.25 | 1 |  |
| U2 | 60s | R | N/A | L | 1 | 1 | 1 |  |
| U3 | 40s | R | N/A | L | 1 | 1 | 1 |  |
| U4 | 60s | R | N/A | L | 1 | 1 | 1 |  |
| U5 | 40s | R | N/A | L | 1 | 1 | 1 |  |
| U6 | 40s | R | N/A | R | 1 | 1 | 1 |  |

Participants of similar age were chosen for both groups to provide some insight on differences between groups and how adults over 40 respond to the stimulation. ‘Unaffected users’ had no reported sensorimotor injury or disease, other than changes with age. We did not use the unaffected side of stroke survivors for comparison because there is known to be some change to the ipsilateral side after stroke [Boll 1974; Kim and Choi-Kwon 1996]. We used the Semmes-Weinstein monofilament exam (SWME) to measure tactile perception on their hand and arm. Participants with mild or moderate sensory dysfunction were chosen because those with severe sensory loss may be frustrated by being unable to sense any signals during the 1.5 hour study.

### 4.2 Survey

Participants took a brief survey after every signal. Our survey contained three parts. The first part contained likert scales for the participant to rate valence and arousal in response to the most recent stimulus. For these questions, numbers 1–9 were associated with pictures of the Self Assessment Manikin (SAM) [Morris 1995].

The next part of the survey allowed participants to select terms to describe how their hand/arm felt in response to the stimulus. The terms were chosen based on pilot testing on users with and without stroke. The options are “pain,” “tickle,” “sensitized,” “tingling/pins and needles,” “numb,” “I can’t feel it,” and “annoying.” Participants were told that they can select one, several, or none of the choices. This list may prime participants, but was a necessary structure. Asking participants to type their feedback is not an accessible solution for either user group. Rapid and standardized response were necessary so that the duration of the study was limited to 1.5 hours to be reasonable for this population. In view of the results – in which all participants selected certain tags far less than others – it is within reason to believe that the list of choices did not solely influenced their response.

Lastly, participants were asked to give verbal feedback on other terms to describe their impression of the signal. The experimenter may then ask follow-up questions. At the end of the study, participants were asked to name their favorite device.

### 4.3 Analysis

Valence and arousal ratings were analyzed using the Wilcoxon rank-sum test (Mann Whitney U test) between two groups, and the Kruskal-Wallis test for comparing more than two groups. Paired t-tests were used to compare average ratings for each of the stimuli between groups. Reported sensations were analyzed between conditions using the chi-squared test. Statistical significance is defined at the *α* = 0.05 level. Verbal comments were analyzed using thematic content analysis. Only three codes emerged from this analysis that were different from the given survey responses: *enjoyable, can barely feel it*, and *strong/intense*.

## 5 Results

### 5.1 Valence and Arousal

Ratings of 1 to 9 were adjusted to –4 to 4 for Figures 4, 5, and 6. Most responses were close to the origin in the high-arousal, positive-valence quadrant, which is associated with terms such as “happy” and “excited.” Users with history of stroke showed similar ratings of arousal as participants in the age-matched unaffected group in response to all stimuli. A paired two-tailed t-test suggests a small difference between the groups’ arousal ratings for each stimuli (t(11) = 2.75, p = 0.02). and their responses are graphed in Figure 4b.

**Fig. 4.**
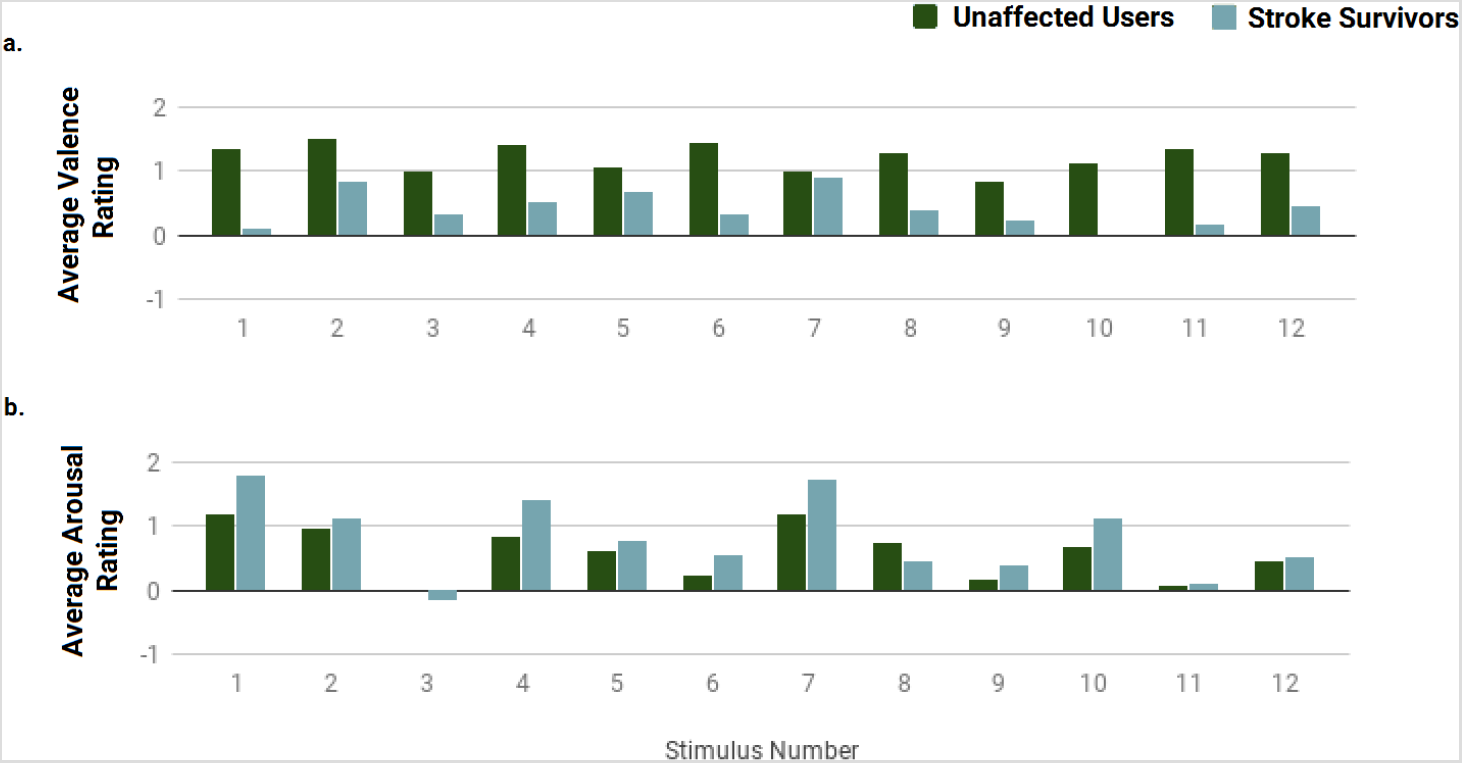
Average valence and arousal ratings for each of the 12 stimuli settings. Ratings here are adjusted to a scale of –4 to 4 (from the 1–9 ranking visible on the survey).

**Fig. 5.**
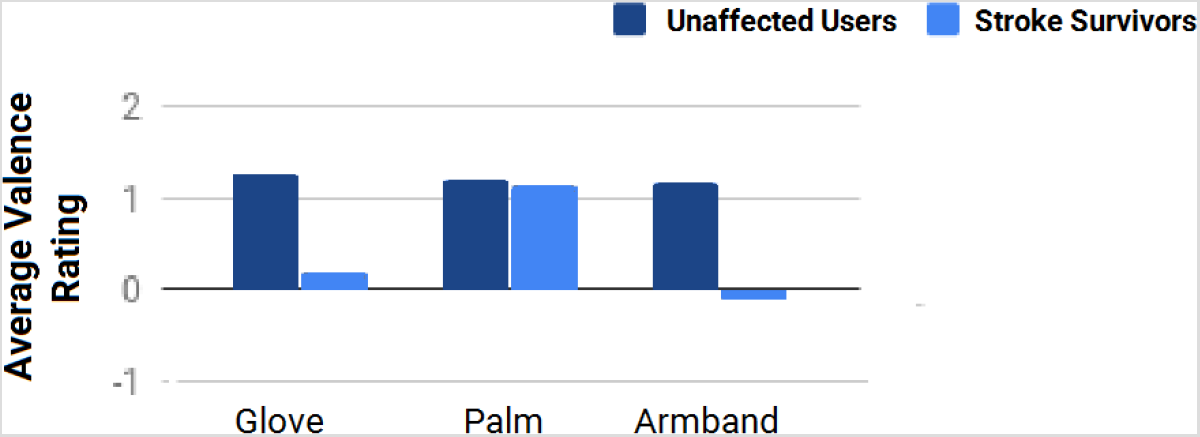
Average valence ratings for stimuli from each device. Stroke survivors report higher valence using the palm stimulation device.

**Fig. 6.**
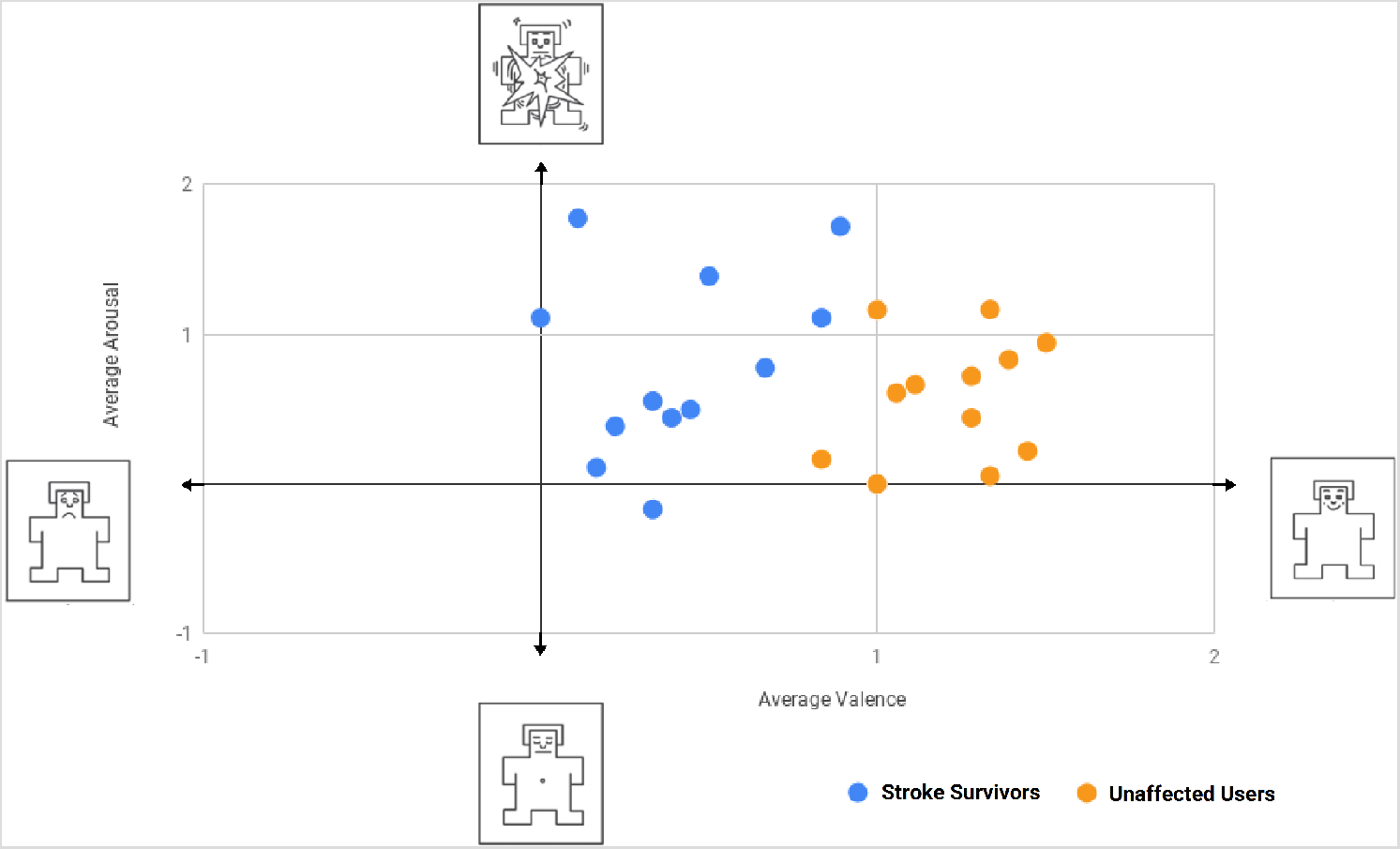
Average valence and arousal rating for each stimulus. Ratings of 1 to 9 were adjusted to –4 to 4. Each dot represents average ratings for one of the twelve stimuli. Icons are adapted from the Self Assessment Manikin (SAM).

Participants responded with the highest arousal ratings in response to the ERM signals, followed by the higher-amplitude LRA signals (significant difference between these factors found using Kruskal-Wallis test H = 15.61, 2 d.f., p = 0.0004). Participants also verbally labeled 71% of ERM stimuli as “strong” (or “intense”), in contrast to 11% or 2% of the high- and low-amplitude LRA signals.

Data show no significant difference in arousal ratings between different activation times or patterns; however, both user groups rated stimuli from the armband device with the lowest arousal ratings. Participants with history of stroke rated the palm device significantly higher arousal than did participants in the unaffected older adults group (Wilcoxon rank-sum test U = 263.5, p< 0.001).

The valence and arousal ratings are plotted together in Figure 6. There is a significant difference in these values between groups. Specifically, Figure 4 shows that while there is only a small difference in arousal ratings between groups, there is a large difference in valence ratings between groups (paired two-tailed t-test t(11) = –8.32, p< 0.00001). Stroke survivors gave consistently lower ratings of valence. The unaffected older adults rated stimuli of lower activation duration (300 ms) with higher valence, but stroke survivors showed no preference between different activation durations.

Both groups rated low-amplitude LRA signals with lowest valence among the motor types and stimuli with a changing location (pseudo-random pattern) with higher valence than stimuli that repeatedly stimulated the same area. Participants also verbally labeled 20% of pseudo-random stimuli as “enjoyable” (“fun,” or “good”).

When comparing valence ratings between devices for all stimuli, stroke survivors rated stimuli from the palm device significantly higher valence than the other devices (Kruskal-Wallis test H = 16.67, 2 d.f., p = 0.00024). The unaffected older adults group did not give significantly different valence ratings to stimuli from different devices (Figure 5).

### 5.2 Reported Sensations

Our first aim was to evaluate if any of the vibrotactile signals were perceived as painful or unacceptable by our participants. None of the signals were reported as painful.

Participants responded that they could not feel or could “barely feel” most stimuli on the forearm, with stroke survivors reporting more frequently that they could not feel the stimulus at all. When participants were asked if they liked that they could not feel the device stimulating, 11/12 participants said “no.” *“If I can’t feel it, I don’t know if it is on,”* said participant U4. However, stimuli from the armband were also the least labeled annoying (chi-squared test *X*^2^(2) = 11.59, *p* = 0.003). Although participants could not feel many stimuli from the armband, they still reported sensations at average rates of 50–80% of the sensations reported from the other devices.

Stimuli on the palm and those using the ERM motors were most labeled ticklish (chi-squared tests suggest the differences are significant: device *X*^2^(2) = 10.82, p = 0.005, actuator *X*^2^(2) = 18.14, p = 0.0001). Participants also reported the ticklish sensation more with stimuli using the pseudo-random pattern, but this difference was not statistically significant at the level of 0.05 (*X*^2^(1) = 3.38, p = 0.066) and was dominated by participants without history of stroke. Figure 7 shows the percentage of stimuli in each category which were labeled ticklish.

**Fig. 7.**
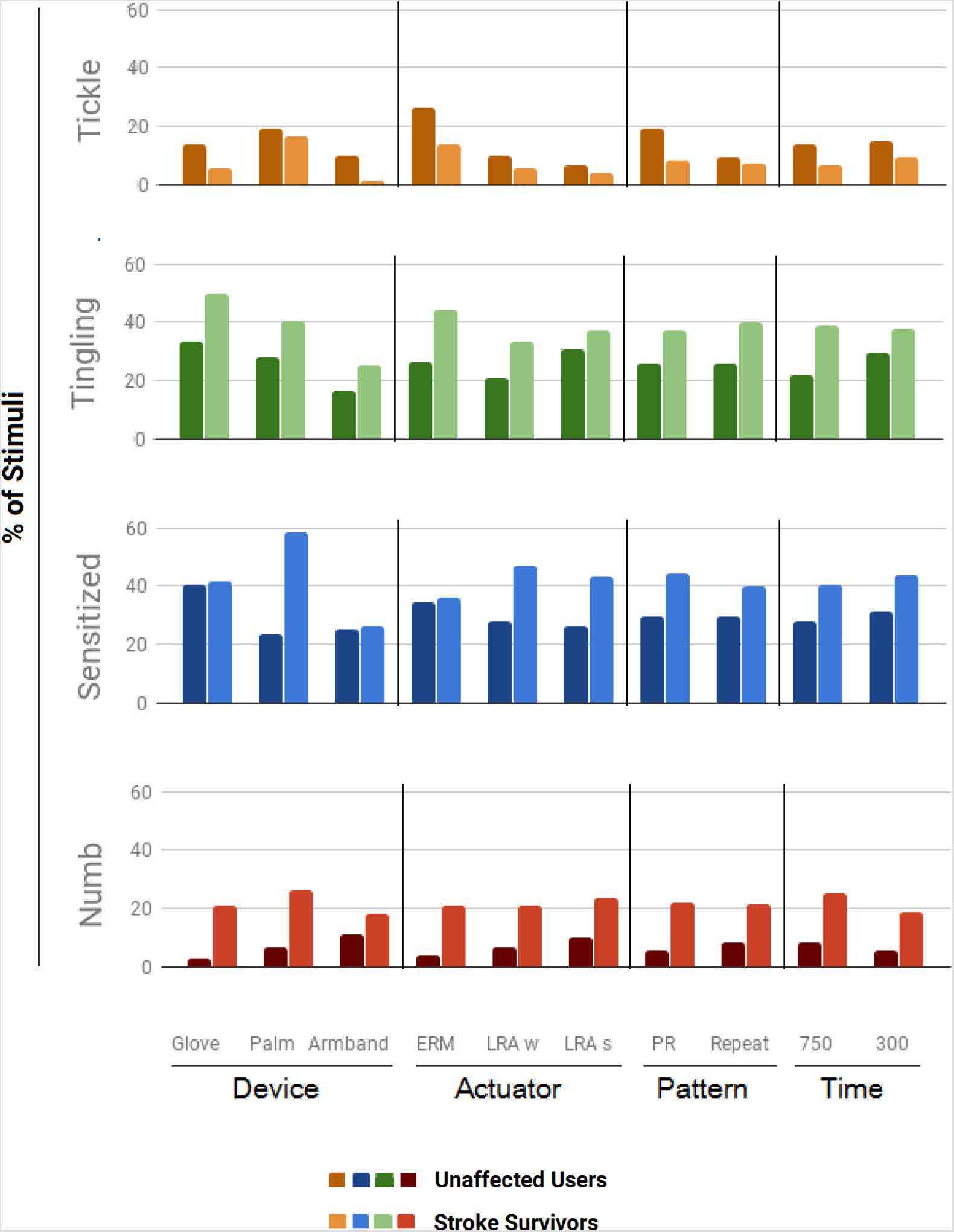
Percent of stimuli in each category that were labeled as giving the hand/arm a tickled, tingling, sensitized or numb sensation.

Stroke survivors report feeling sensitized, tingling, or numb at higher rates (150–200%) across all stimuli than participants in the unaffected older adults group. This group also reported feeling sensitized almost three times the rate of the unaffected group when using the palm device. Phalanges stimulation was most associated with tingling out of the three locations (a chi-squared test suggests the difference is significant: *X*^2^(2) = 14.66, p = 0.0007).

When asked about which device/stimulation zone that they preferred, the phalanges-stimulating glove was chosen by 6 (50%) of participants in both groups. Only one participant chose the armband device, and 5 chose the palm stimulation device. Participants are not informed of the potential applications for this stimulation, however, they expressed preferences and beliefs. Four participants expressed a belief that strong stimulation is better. When discussing why, they said:

*“I want [the stimulation] even stronger. I like deep tissue massage.”* – U2

*“I have a TENS [electrical stimulation] unit, and eventually I turned it all the way up. So I expect, that I may want to adjust this all the way up.”* – S1

*“It is like TENS [electrical stimulation]. I like to use mine at the highest setting. So that is why I want strong.”* – S3 *“When you work out, you don’t just la-de-da work out. You work out strong.”* – S6

## 6 Discussion

### 6.1 Valence and Arousal

Data suggest that stimuli from the armband are found to be less arousing than stimuli applied to the hand. Stimuli with lower amplitude are also rated as less arousing. Participants report that they can barely feel stimuli from the armband, as would be true with very low amplitude stimuli, so intensity of sensation may be a factor in arousal as found in prior work [Hasegawa et al. 2019; Yoo et al. 2015]. Participants also expressed dissatisfaction with these signals, so higher arousal stimulation may be acceptable when compared to other factors.

The disparity in valence ratings between groups may be due in part to different preferences for activation duration. Two participants in the stroke survivors group described difficulty sensing time differences in the signals. Some survivors may have increased latency to sense their affected side after stroke [Boivie et al. 1989; Peurala et al. 2002]. Stroke survivors may have difficulty sensing temporal differences in stimuli, which may impact their impression of and preference for vibrotactile signals.

*“I can kind of sense that there is a pattern or timing to the signals, but I can’t feel what it is. I can only tell whether it is moving around or is in one place and how strong it is. I am slow to sense it or something, so I can’t tell any more than that.”* – S6

### 6.2 Reported Sensations

Sensations felt in the hand and arm may help us understand the body’s response to stimulation and inform interaction designers. Participants in both groups reported that they could not feel or could barely feel most signals applied to their forearm. This is likely because the density of cutaneous mechanoreceptors is much less in the arm compared to the fingers and hand [Chambers et al. 1972; Guyton 1996; Johansson and Vallbo 1979; Lee 2010]. It is worth noting that the muscle afferent fibers, which sense mechanical stimulation of the muscles, are also present and provide sensory feedback to the central nervous system like the mechanoreceptors [Johansson and Vallbo 1979]. These sensory receptors are located in the muscle belly. The forearm contains more muscle groups than the hand. One may predict that subthreshold stimulation is desirable, however participants did not like stimuli they could not feel.

Participants expressed more tickle feeling in response to palm stimulation, perhaps because this is a more ticklish area of the body than the top of the fingers or the forearm [Harris 1999]. Participants also expressed increased tickle feeling in response to changing (pseudo-random) and erratic mechanical stimuli (ERM motors), possibly relating to how expected or predictable stimuli are less ticklish [Harris and Christenfeld 1999].

Participants often reported a tingling sensation during and after stimulation, particularly when using the hand stimulation devices. Tingling can be associated with hand-arm vibration (HAV) syndrome or “vibration white finger” – a condition found in practitioners such as dentists and construction workers, who use tools such as drills and jackhammers for extended periods of time. The condition begins to set in after consistent exposure to high-amplitude vibration for more than 6 months. Stimuli here are far below the ISO 5349 standard limits of exposure [Bovenzi 1998], so tingling here most likely relates to the sensitizing or analgesic effects of vibration. Vibrotactile stimulation is used to reduce pain associated with injections, teething and chronic conditions [Guieu et al. 1991; Hagura et al. 2013]. Side affects such as tingling are reported during such uses [Sharma et al. 2011].

This study has limitations primarily regarding sample size and duration of testing; however, this work provides key preliminary insights on user impressions and beliefs.

## 7 Conclusion

Vibrotactile stimulation is being used in a number of contexts, with emerging application to older adults and survivors of brain injury. To inform the development of more user-conscious systems, here we present a results of a study on older users’ impressions of vibrotactile stimuli.

None of the stimuli settings were found to be intolerable or painful. Low-amplitude stimuli were found to be the least arousing. Stroke survivors report higher rates of some sensations but lower valence overall than those without history of stroke. Participants expressed a preference for stimulation with a changing activation site (pseudo-random spatial pattern), calling these stimuli more “enjoyable.” Forearm stimulation was the least preferred here, although these stimuli were the least arousing between devices. Participants in both groups (with and without history of stroke) expressed dissatisfaction with stimulation which they could not sense. Similarly, a third of participants explicitly requested the strongest (high-amplitude) stimuli, likening it to massage or TENS. Participants also provided feedback on what sensations they felt in their hand and arm. Reported sensations include tickle, tingling, and numbness. User preferences such as these can be considered along with physiological, safety, and hardware constraints in the development of interactive technologies.

## Data Availability

Not Applicable

## 8 Acknowledgments

This research was supported, in part, by the Stanford Wu Tsai Neurosciences Institute Neuroscience:Translate Program and Postdoctoral Fellowship #1F32HD100104–01 from the National Institutes of Health (NIH).

## Footnotes

1 Stroke is an acquired brain injury.

2 Laboratory measurements were repeated for confirmation. At this frequency, the motors are expected to have an amplitude of less than 1 g. Measure actuator performance for your given setup.

